# Online media reporting of global mortality rates and causes: a content analysis

**DOI:** 10.1101/2025.04.22.25326092

**Authors:** Trevor Treharne, Carl Heneghan

## Abstract

**Introduction:** The online news media covers mortality-related topics such as illnesses, diseases, violent deaths, and fatalities caused by environmental or lifestyle factors. This frequent coverage, coupled with the media’s significant influence on public opinion, can make the public overestimate certain health risks while underestimating others.

**Objective:** This study aggregates global death figures from online media reports, compares them to official annual death figures, and analyses online media’s over- or under-reporting of specific death causes.

**Design:** This content analysis examined online media articles from 1 January 2021 to 31 December 2021, citing mortality figures for different causes of death. By utilising global news databases Dow Jones Factiva and Nexis and the news aggregator service Google News, it identified articles that referenced annual death statistics for various causes. These findings were used to determine the overall global death count based on aggregated figures from online media reports on causes of death. A comparison was then made with the official global death total. Additionally, each cause of death was compared to the most reliable secondary source, such as the best available official figures or academic research, to discern areas where the media may exaggerate or downplay specific causes.

**Results:** The media attributed more than twice the actual deaths in 2020, RR =2.43: 155 million media compared (20.1 per 1000) to the UN-reported 63.7 million deaths (8.3 per 1000). Official or academic sources also overestimate the number of deaths, with 144.3 million deaths reported annually (18.7 per 1000). Across 44 death-cause categories, the media’s figure for annual deaths matched the best available source ten times (23%) but failed to identify the best source in the other 34 categories. Notable areas where the media over-reported causes included Influenza (65,000 reported vs 14,500 actual, RR:4.48), natural disasters (60,000 reported vs 15,071 actual, RR:3.97) and overwork (2.8m reported vs 745,000 actual, RR:3.76).

**Conclusions:** Online media reports indicate an overestimation of global mortality compared to the actual annual mortality rate and the rate supported by official and academic research sources. There is a tendency for over-reporting of death rates in areas that attract media attention, such as influenza and natural disasters, while there is under-reporting in less attention-grabbing areas, like digestive and liver diseases.

## Introduction

While the media are highly influential in shaping discourses about health (Wallack & Dorfman, 2001; Hayes et al., 2007), the relationship between media coverage of health issues and their tangible public health risk can become disparate, with an inverse relationship between the frequency of media reports and the number of fatalities for the health risks covered (Bomlitz & Brezis, 2008).

The news media has a persistent focus on producing reports about death (Hanusch, 2010). Of CNN’s top 100 digital stories of 2021, 24 were related to death Andrew et al., 2022). Of Upday News’ ‘20 biggest news stories in UK media in 2021’, seven were associated with death (King, 2021).

The ‘Perils of Perception’ 2020 survey by Ipsos asked members of the public in 32 nations for the leading causes of death in their country. It found that people underestimate the two main causes of death – cardiovascular diseases and cancer – and overestimate more infrequent death causes such as accidental deaths, transport injuries, suicide, interpersonal violence, substance abuse and terrorism (Ipsos, 2020). The Ipsos report cites media coverage as a likely factor influencing these findings.

Media reporting can directly impact the mental state of readers and viewers. For example, the emotional toll of watching negative television news bulletins and programmes was measured (Johnston & Davey, 1997) and demonstrated increased anxiety and unhappiness after just 14 minutes of viewing.

During the COVID-19 pandemic, there was recurrent coverage of the disease, with 28 million online news media stories related to COVID in 20 countries from October 2019 to May 2020 (Ng et al., 2021). The consequence of such coverage is that “endless newsfeeds related to COVID-19 infection and death rates could considerably increase the risk of mental health problems” (Suet et al., 2021). Furthermore, a positive correlation between media coverage of COVID-19 and the “severity of unspecific anxiety, depression and topic-specific fear” has also been discovered (Bendau et al., 2021).

Increased and disproportional public reactions to highly infrequent death causes, such as shark attacks, have also been linked to the media’s approach to their coverage(Eovaldi et al., 2016). Death anxiety, sometimes termed as ‘thanatophobia’, is considered “omnipresent”, “affects each and every one of us” (Sinoff, 2017), and is a “normal human experience”, but if persistent or reoccurring can lead to various psychological conditions (Iverach et al., 2014). Death anxiety can impact our prejudices (Greenberg et al., 1990) and even influence which politicians we vote for (Cohen et al., 2004).

Considering the media can impact public health behaviour – including influencing vaccination rates (Begg et al., 1998), health services utilisation (Grilli et al., 2002), diagnostic testing demands (Sharma et al., 2003) and prescription drug compliance (Matthews et al., 2016) – its framing and reporting of death causes and overall rates is likely to prove influential.

According to the United Nations’ World Population Prospects, 63.7 million people died globally in 2020 (United Nations, 2022) from a global population of 7.7 billion (United Nations, 2019) meaning 0.82% of the world’s population suffered death. The media provides broad coverage of death and health stories, ranging from violent attacks and tragic accidents to common diseases and rare illnesses.

A common strand of media reporting is to use global statistics of the disease or death cause in question to provide editorial emphasis. These stats are most frequently cited in isolation and not in the context of the 63.7 million people that die globally.

This study aimed to establish a global death figure based on online media reports and compare it to the UN’s figure and then a reliable secondary source. By examining major death causes, we sought to analyse the specific causes that are overrepresented or underrepresented in online media coverage.

## Methodology

### Sample

This study collected articles from international newspapers and dedicated news sites in English using three sources: global news databases Dow Jones Factiva and Nexis, which provide access to numerous online news sources worldwide, and Google News, a news aggregator service developed by Google. The articles analysed were published between 1 January 2021 and 31 December 2021. Excluded from the analysis were articles from specialised health and medicine websites, non-news sites like WHO or charity websites, and academic sources such as The Lancet, BMJ, and other journals.

### Search methods

For global news databases Dow Jones Factiva and Nexis, only the ‘All Publications’ OR ‘All Web News’ sections were used. The search terms “deaths a year,” OR “annual deaths”, OR “deaths per year”, OR “cause of death”, OR “global mortality” were used. This returned 8,359 results. For both Dow Jones Factiva and Nexis, the headlines and introductory text were scanned for relevant articles reporting on global death causes. Death causes were then searched for using the Institute for Health Metrics and Evaluation (IHME) Global Burden of Disease report, which lists the 31 leading causes of death contributing to at least 0.1% of annual global deaths. These causes of death were searched on Factiva, Nexis and Google News.

### Data collection

Data collected was the article title, publication, publication country, publication date, link to article, cause of death, deaths per annum (according to the article), and article source (where the news article found the death figure). Then a second source was sought to check each death category and the number of deaths for that cause in official statistics. This second source comparison focused on finding the best available evidence for each death category. When there is more than one suitable media or second source death rate figure, the higher figure is chosen to ensure consistency.

### Data analysis

There was an overarching analysis of the total number of global deaths according to an aggregate figure of online media reports compared to a second official or academic source’s total number of global deaths. Further, there was a comparison between the number of fatalities reported for a specific death in the media and the second source, enabling an analysis of which specific death causes the media exaggerates or underplays.

## Results

While 63.7 million people died from a global population of 7.7 billion (8.27 per 1000) in 2020, the media attributed 155 million annual deaths to 44 major death causes (20.1 per 1000). This means that media reporting of annual deaths is 2.43 times higher than the yearly death rate. The overall death figure from the second source found there are 144.3 million deaths annually (18.7 per 1000), 10.7 m less than the media figure.

Of the 44 death cause categories, the media’s figure for annual deaths matched the best available source ten times but failed to identify the best source in the other 34 categories.

**Table 1:**
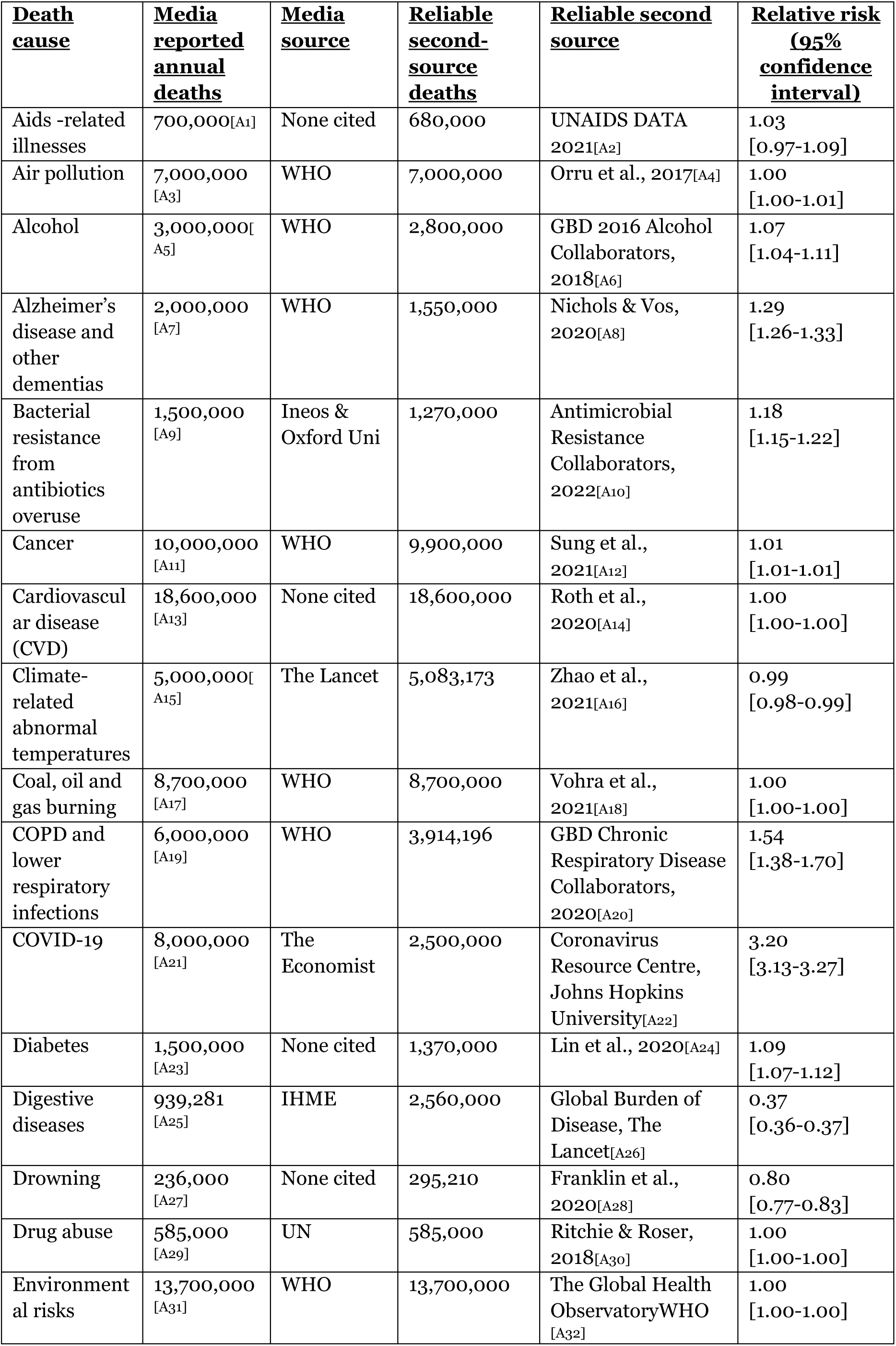
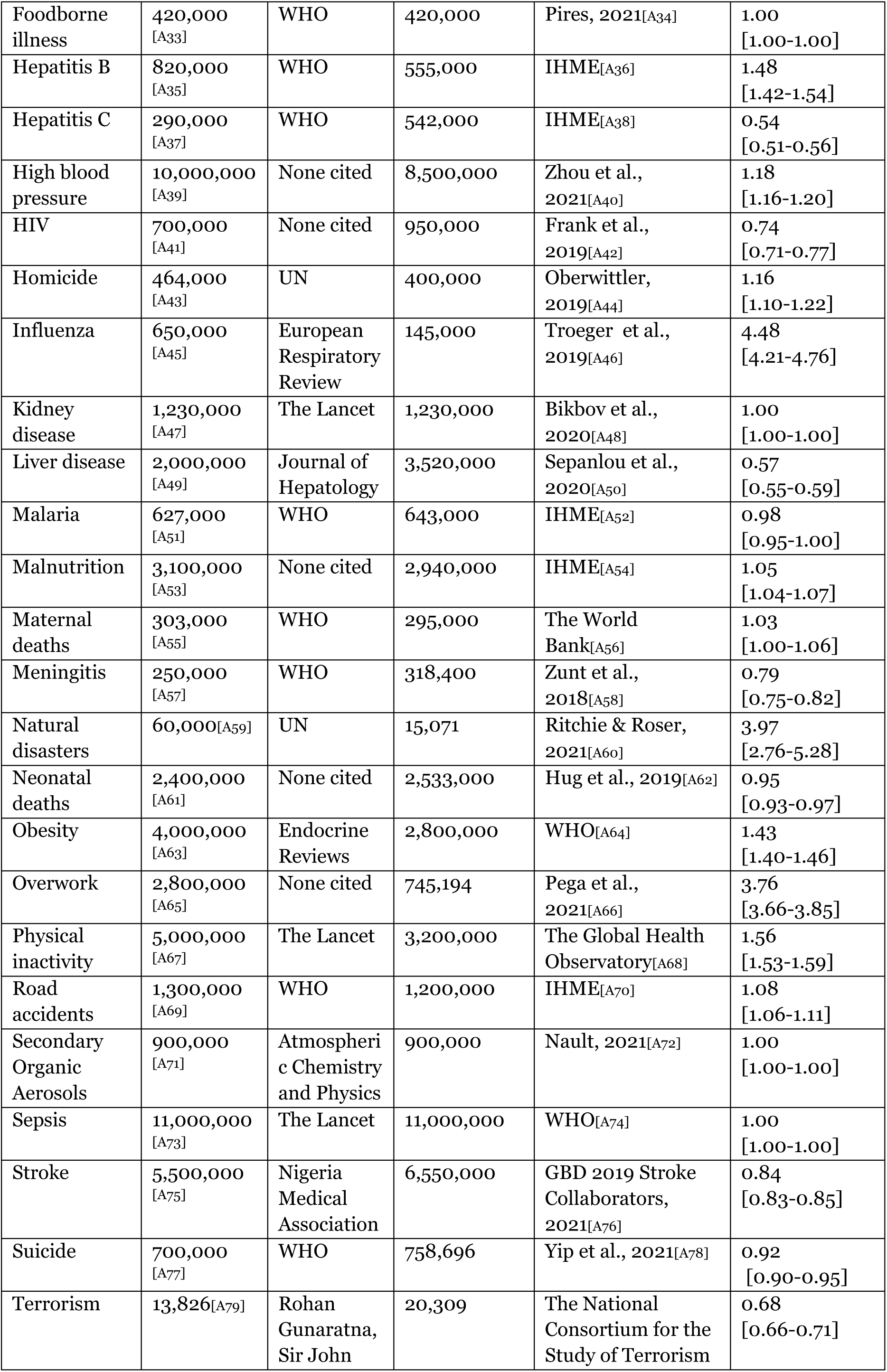
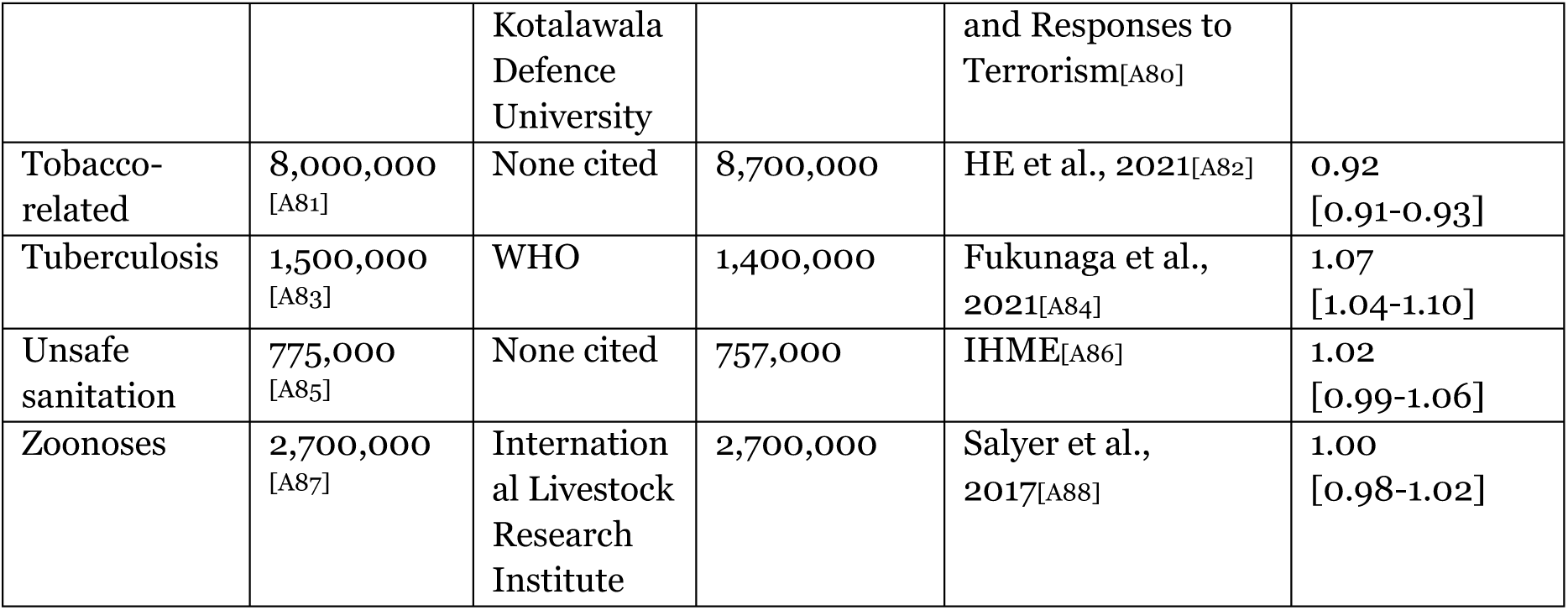
Major death causes by annual deaths – media reported and reliable second source. *Media reporting of death causes and second source death cause statistics*

Figures 1 and 2 show those causes where the media substantially over- or under- reported specific deaths. Deaths were over-reported for Influenza (65,000 reported vs 14,500 actual, RR:4.480), Natural disasters (60,000 reported vs 15,071 actual, RR:3.970), and Overwork (2.8m reported vs 745,000 actual, RR:3.760). Under-reported deaths included digestive diseases (93,000 reported vs 256,000 actual, RR:0.370), Hepatitis C (290,000 reported vs 542,000, RR:0.540), and liver disease (2m reported vs 3.5m actual, RR:0.570).

**Figure 1:** The relative risk ratio of the media-reported annual deaths and the reliable second source annual deaths (death causes at the top represented the most considerable media exaggeration, those at the bottom the biggest under-representation) [In supplementary material – Slide 1]

**Figure 2:** The percentage difference between the media-reported annual deaths and the second source annual deaths (death causes at the top represented the most considerable media exaggeration, those at the bottom the biggest under-representation) [In supplementary material slides – Slide 2]

## Discussion

This study suggests that online media reports consistently depict a higher global mortality rate than the actual annual mortality rate and official research sources. The media tends to overstate death figures, reporting more than double the number of deaths documented by the United Nations. Additionally, the study identifies specific instances of over- and under-reporting by the media, highlighting areas such as influenza, natural disasters, and overwork as prominent examples.

These findings underscore the importance of exercising caution and critical assessment when consuming online media reports on mortality rates and causes. It becomes evident that certain causes of death, particularly those that attract attention and sensationalism, are more susceptible to over-reporting, while other causes, like digestive and liver diseases, may be under-reported due to their comparatively lower media appeal.

This problem likely occurs because journalists often want more tentative but fresh or dramatic medical research to cover (Nelkin, 1996). When considering if a story is newsworthy, health journalists often consider if the topic is “something that affected audiences directly” (Leask et al., 2010). This results in personal health and wellbeing, alongside common conditions, proving especially popular in media coverage. In this study’s breakdown of over and under-reporting of specific death causes, there is a greater exaggeration for such widely reported topics. Of the most over-reported death causes, several are aligned with those most likely to affect audiences and, as such, be more appealing as a media story. For example, influenza and natural disasters are popular media topics and attract the most death cause exaggeration.

Overwork is a prevalent topic as media audiences are concerned by the impact of hectic work schedules. Other over-reported death causes – such as COVID, physical inactivity, obesity, homicide and road accidents – align with how the media view an appealing story for their audiences.

Alternatively, the most under-reported death causes are often unappealing or niche areas, such as digestive diseases, Hepatitis C and liver disease. Further, a death cause identified to cause copycat incidents, suicide, is underplayed by the media, most likely for ethical reasons (Etzersdorfer & Sonneck, 1998).

This implies that editorial agendas significantly emphasise death reporting on popular topics among journalists, media outlets, and their readers while inversely underreporting less popular areas. As a result, the portrayal of the death cause landscape may become skewed, shaped by the media’s coverage agenda rather than actual evidence on causes of death.

Furthermore, the number will be higher because of the presence of comorbidities (Valderas et al., 2009). For example, 27% of US adults have multiple chronic conditions (Boersma at al., 2020); in the UK, 54% of people over 65 have two or more conditions, and 17% of middle-aged and older people in China have two or more chronic diseases simultaneously (Fan et al., 2021).

During the COVID-19 pandemic, people with comorbidities were particularly susceptible to adverse outcomes, including death (Clark et al., 2020). In addition, comorbidities make it more difficult to precisely assign the cause of death, leaving reporting of death data with accuracy issues in assigning causation (Jefferson et al., 2020).

The media’s reporting of specific deaths lacks contextualisation regarding comorbidities and other contributing factors associated with those causes. Typically, the media presents death causes in isolation, emphasising singular gross figures.

Consequently, consumers of media are left with the perception of a diverse range of death causes, all linked to a significant annual death figure attributed to each cause. This creates an overarching impression of pervasive and multifaceted threats to life on a large scale, despite the likelihood that most of these causes will never directly impact the individual media consumer.

To enhance the current state of reporting, the media should provide contextual information alongside individual death cause figures. For instance, instead of presenting statements like “Raised blood pressure is the biggest cause of death globally, *causing more than* 10 million deaths per year” (*The Independent***Error! Bookmark not defined.**), a more aligned approach to source terminology could be adopted, such as “8.5 million deaths *were associated with* high blood pressure” (*Nature Reviews Cardiology***Error! Bookmark not defined.**). This distinction is crucial, as an association should not be conflated with causality (Altman & Krzywinski, 2015). While causation necessitates an association between variables, the presence of an association does not necessarily imply causation (Zhang & VanDyke, 2022). By employing a linguistic shift from ‘cause’ to ‘association’, media reports could elicit a less alarmist reaction among media consumers when encountering information about a specific death cause.

Media consumers should critically evaluate and adopt a sceptical mindset when consuming online media reports on mortality rates and causes. It is essential to take into account the underlying motives and potential sensationalism that can shape media coverage. By exercising critical thinking and seeking out additional reliable sources, readers can better understand global mortality trends and the specific causes of death.

To obtain dependable information on global mortality rates and causes, readers should turn to authoritative sources such as the Office for National Statistics (ONS), the United Nations (UN), the World Health Organization (WHO), the Centers for Disease Control and Prevention (CDC), and the Global Burden of Disease (GBD) study conducted by the Institute for Health Metrics and Evaluation (IHME).

## Limitations

The study considers online global media as a heterogeneous information source. However, international media is fragmented, and various geographies approach content differently. While all sources analysed in this research are major international media outlets, their widespread distribution poses challenges in forming a specific regional or national perspective.

The complex impact of the COVID-19 pandemic on global death rates and comorbidities likely influences the study’s findings. The precise effects of COVID-19 on mortality rates have been challenging to ascertain and may have influenced the outcomes of this study.

The findings of this study may vary across different years due to potential shifts in media reporting practices, changes in public health priorities, or fluctuations in global mortality rates. Therefore, independent replication of this research is crucial to validate and corroborate the findings over different periods.

It is vital to acknowledge that the results of this study may exhibit variations across countries due to differences in media landscapes, cultural factors, and regional disparities in mortality reporting.

Consequently, the applicability and generalisability of the findings at the country-specific level may be limited, highlighting the necessity for further research encompassing diverse geographical contexts to obtain a more comprehensive understanding of how the media portrays global mortality rates and causes.

## Conclusion

The media’s coverage of death and specific causes substantially influences public perceptions of mortality and likely causes of death. The present study reveals that online media reports consistently depict a higher global mortality rate than the actual annual mortality rate and official/academic sources. Notably, there is a tendency to over-report death rates in areas that attract media attention, while under-reporting prevails in less appealing areas.

## Data Availability

All data produced in the present study are available upon reasonable request to the authors

